# Functional and quantitative evaluation of BNT162b2 SARS-CoV-2 vaccine-induced immunity

**DOI:** 10.1101/2021.05.26.21257884

**Authors:** Hitoshi Kawasuji, Yoshitomo Morinaga, Hideki Tani, Yumiko Saga, Makito Kaneda, Yushi Murai, Akitoshi Ueno, Yuki Miyajima, Yasutaka Fukui, Kentaro Nagaoka, Chikako Ono, Yoshiharu Matsuura, Hideki Niimi, Yoshihiro Yamamoto

## Abstract

**Objectives:** Vaccines against severe acute respiratory syndrome coronavirus-2 have been introduced. To investigate whether the vaccine provides protective immunity effectively, the amount and function of vaccine-induced antibodies were evaluated.

**Methods:** Sera from 13-17 days after the second dose of the Pfizer BNT162b2 vaccine were collected from healthcare workers at the University of Toyama (n=740). Antibody levels were quantitatively measured by the anti-receptor binding domain antibody test (anti-RBD test), and neutralising activity against pseudotyped viruses displaying wild-type (WT) and mutant spike proteins (B.1.1.7- and B.1.351-derived variants) were assayed using a high-throughput chemiluminescent reduction neutralising test (htCRNT). Basic clinical characteristics were obtained from questionnaires.

**Results:** Antibodies were confirmed in all participants in both the anti-RBD test (median 2112 U/mL, interquartile range [IQR] 1275–3390 U/mL) and the htCRNT against WT (median % inhibition >99.9, IQR >99.9 to >99.9). For randomly selected sera (n=61), 100.0% were positive for htCRNT against the B.1.1.7- and B.1.351-derived variants. Among those who answered the questionnaire (n=237), the values of the anti-RBD test were negatively correlated with age for females (p<0.01; r = -0.31, 95% confidence interval -0.45 to -0.16). Systemic symptoms after vaccination were related to higher values of the anti-RBD test (median 2425, IQR 1450 – 3933 vs. median 1347, IQR 818 – 2125 for no symptoms; p<0.01).

**Conclusions:** The BNT162b2 vaccine produced sufficient antibodies in terms of quality and quantity which could neutralise emerging variants. Antibody induction can be affected by age and sex but will still be at a sufficient level.

## Introduction

Since severe acute respiratory syndrome coronavirus-2 (SARS-CoV-2) appeared in China, the COVID-19 pandemic has strongly limited our healthy living and economic activity. There are still no therapeutic breakthroughs, but vaccination is expected to be a specific and effective defence strategy to fight COVID-19. Several vaccines against SARS-CoV-2 have been introduced, and to date, vaccines manufactured by Pfizer (BNT162b2) have been administered in Japan. This vaccine was shown to have preventive effects in 95% of individuals against COVID-19 [1]; however, SARS-CoV-2 variants have emerged after the report.

The B.1.1.7 variant, which originated in the United Kingdom, has a mutation (N501Y) related to the increased affinity of the virus to the ACE2 receptor expressed on targeted cells [2]. The B.1.351 variant, which originated in South Africa, has mutations K417N, E484K, and N501Y in the receptor binding domain (RBD) of the spike (S) protein, which are related to reduced neutralisation [3]. The effectiveness of the Pfizer vaccine against these emerging variants has been reported to be similar to that of wild-type SARS-CoV-2 [4]. In contrast, a reduction in the neutralisation potential of vaccinated sera against variants has also been reported [5-7]. Therefore, a continuous evaluation of vaccine efficacy is required.

Neutralising antibodies have protective functions against pathogens. We previously established the chemiluminescence reduction neutralization test (CRNT) for the evaluation of neutralising activity against SARS-CoV-2 using pseudotyped virus [8]. The high-throughput CRNT (htCRNT), which is a modified method using 384-well microplates, also has a good correlation with the CRNT [8]. These methods assess the inhibition of infectivity in target cells. In contrast, it is impossible to assess protective function directly by commercially available antibody tests because they do not assess inhibition against viral infection. However, some tests can provide quantitative values of antibodies and help us to speculate on the state of acquired immunity. In this study, we confirmed that the Elecsys Anti-SARS-CoV-2 S immunoassay, which is an anti-RBD antibody test, and our neutralising tests are well-correlated and can effectively identify convalescent COVID-19. These findings suggest that it is possible to quantitatively and qualitatively evaluate antibodies against SARS-CoV-2 using these two tests.

In confronting emerging variants, it is necessary to understand the immunity acquired by the BNT162b2 vaccine. In the present study, sera after mass inoculation of healthcare workers were systematically obtained at our institute. Using two antibody assays, one quantitative and the other functional, the robustness and vulnerability of the immune status acquired by vaccination were investigated.

## Materials and Methods

### Collection of specimens

Serum samples were collected from healthcare workers at the Toyama University Hospital. All patients received two doses of the Pfizer BNT162b2 vaccine, and blood was drawn between 13-17 days after the second dose. The sera were used for serological assays on the day of blood collection or within 3 days of storage at 4°C. The remaining sera were frozen at -80°C until further verification.

Basic clinical characteristics were arbitrarily obtained from questionnaires. The following characteristics were obtained: age, sex, local symptoms after vaccination (pain at injection site, redness, swelling, hardness, local muscle pain, feeling of warmth, itching, and others), systemic symptoms after vaccination (fever ≥37.5°C, general fatigue, headache, nasal discharge, abdominal pain, nausea, diarrhoea, myalgia, joint pain, swelling of the lips and face, hives, cough, and others), underlying diseases (malignant diseases, hypertension, diabetes, dyslipidaemia, renal failure, liver failure, asthma, autoimmune diseases, and others), and medications (corticosteroids excluding ointment, immunosuppressants, anti-tumour drugs, anti-rheumatoid drugs, and radiological therapy).

### Generation of pseudotyped viruses

Pseudotyped vesicular stomatitis virus (VSV) bearing SARS-CoV-2 S protein was generated as previously described [8, 9]. The expression plasmid for the truncated S protein of SARS-CoV-2, pCAG-SARS-CoV-2 S (Wuhan), was kindly provided by Dr. Shuetsu Fukushi, National Institute of Infectious Diseases, Japan. The expression plasmids for the truncated mutant S protein of SARS-CoV-2, pCAGG-pm3-SARS2-Shu-d19-B1.1.7 (B.1.1.7-derived variant), and pCAGG-pm3-SARS2-Shu-d19-B1.351 (B.1.351-derived variant), were constructed by PCR-based site-directed mutagenesis using cDNA as a template, which was obtained by chemical synthesis with optimisation for the humanised codon (Thermo Fisher Scientific, Waltham, MA). S cDNA of SARS-CoV-2 was cloned into the pCAGGS-pm3 expression vector. Briefly, 293T cells were transfected with the above expression vectors. After 24 h of incubation, the transfected cells were infected with G-complemented (*G) VSVΔG/Luc (*G-VSVΔG/Luc) at a multiplicity of infection of 0.5. The virus was adsorbed and extensively washed four times with Dulbecco’s modified Eagle’s medium (DMEM) supplemented with 10% foetal bovine serum (FBS). After 24 h of incubation, the culture supernatants containing pseudotyped VSVs were centrifuged to remove cell debris and stored at −80°C until further use.

### Serological tests

The neutralising effects of samples against pseudotyped viruses were examined in 384-well microplates (Corning Inc., Corning, NY) using the htCRNT, as previously described. In this study, we utilized VeroE6/TMPRSS2 cells, which are highly susceptible to SARS-CoV-2 infection. VeroE6/TMPRSS2 cells (JCRB1819) were purchased from the Japanese Collection of Research Bioresources (JCRB) Cell Bank (Osaka, Japan). Briefly, 100-fold diluted serum-containing DMEM (Nacalai Tesque, Inc., Kyoto, Japan) with 10% heat-inactivated FBS was incubated with pseudotyped SARS-CoV-2 for 1 h. After incubation, VeroE6/TMPRSS2 cells were treated with DMEM-containing serum and pseudotyped virus. The infectivity of the pseudotyped viruses was determined by measuring the luciferase activity after 24 h of incubation at 37°C. A score with ≥50% inhibition of viral infection was considered positive for neutralisation.

For the commercial assay, serum samples were tested at Toyama University Hospital using the Elecsys Anti-SARS-CoV-2 S immunoassay (Roche Diagnostics GmbH, Basel, Switzerland) to quantitatively measure antibody levels against SARS-CoV-2 RBD. The manufacturer’s cut-off value (COV) was 0.8 U/mL and the minimum value was expressed as <0.4 U/mL.

### Statistical analysis

Statistical analysis was performed using the Mann-Whitney test to compare non-parametric groups. The Friedman test with Dunn’s test was used for multiple comparisons among the three paired groups. Correlations between test findings were expressed using Pearson’s correlation coefficients. Data were analysed using GraphPad Prism version 8.4.3 (GraphPad Software, San Diego, CA). Statistical significance between different groups was defined as P <0.05. Data are expressed as medians with interquartile ranges (IQR).

### Ethics approval

This study was performed in accordance with the Declaration of Helsinki and was approved by the ethical review board of the University of Toyama (approval No.: R2019167). Written informed consent was obtained from all participants.

## Results

### Antibody quantification and neutralising activity after vaccination

Serum samples were obtained from a total of 740 participants. All participants were positive for the htCRNT and anti-RBD antibody tests (**Fig 1**). All htCRNT values were over 80.0% (range, 80.6-100.0), and the median was >99.9% (IQR >99.9 to >99.9). For the anti-RBD antibody test, the range and median were 28.5-10824 U/mL and 2112 U/mL (IQR 1275–3390 U/mL), respectively.

**Figure 1.**
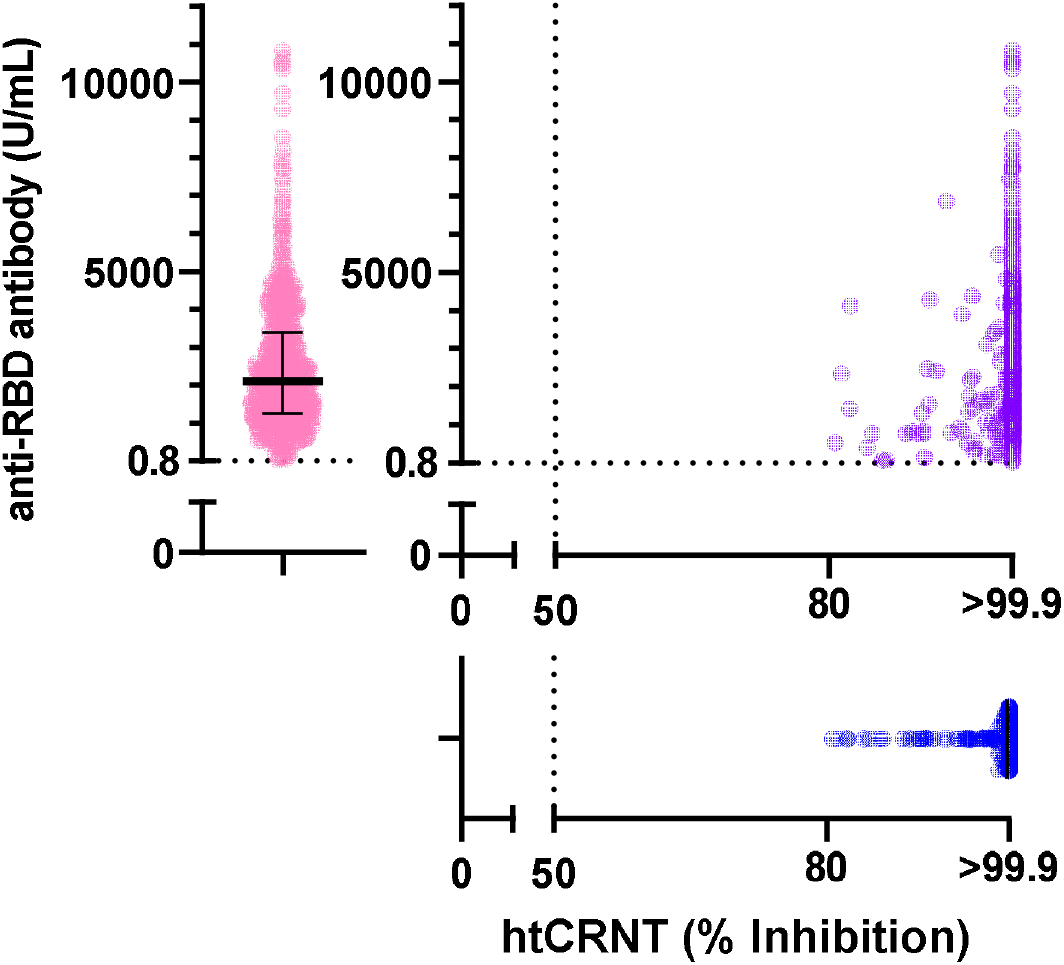
Relationship between anti-RBD antibody levels and neutralisation. Neutralisation levels against pseudotyped viruses measured by htCRNT (blue) and anti-RBD antibody levels measured by a commercially available test (pink) were plotted (n=740). The results of both tests are plotted on the XY coordinate (purple). The value of htCRNT is the mean of duplicate assays using 100-fold diluted serum. Bars indicate medians with interquartile ranges. htCRNT, high-throughput chemiluminescent reduction neutralising test; RBD, receptor-binding domain.

Next, 61 representative samples were randomly selected and assayed using pseudotyped B.1.1.7 and B.1.351-derived variants. Of these, 100.0% (61/61) were positive for htCRNT against both variants. Compared to the wild-type (median >99.9; IQR >99.9 to >99.9), the htCRNT values against the B.1.1.7 and B.1.351-derived variants were significantly decreased (median 97.8, IQR 90.9 - >99.9 and median 96.9, IQR 89.0 - 99.2, respectively; p<0.01) (**Fig 2A**). The values of the htCRNT against the B.1.1.7 and B.1.351-derived variants were positively correlated with the values of the anti-RBD antibody test (r = 0.44, 95% confidence interval [CI] 0.21-0.62 and r = 0.40, 95% CI 0.17-0.60, respectively) (**Fig 2B**).

**Figure 2.**
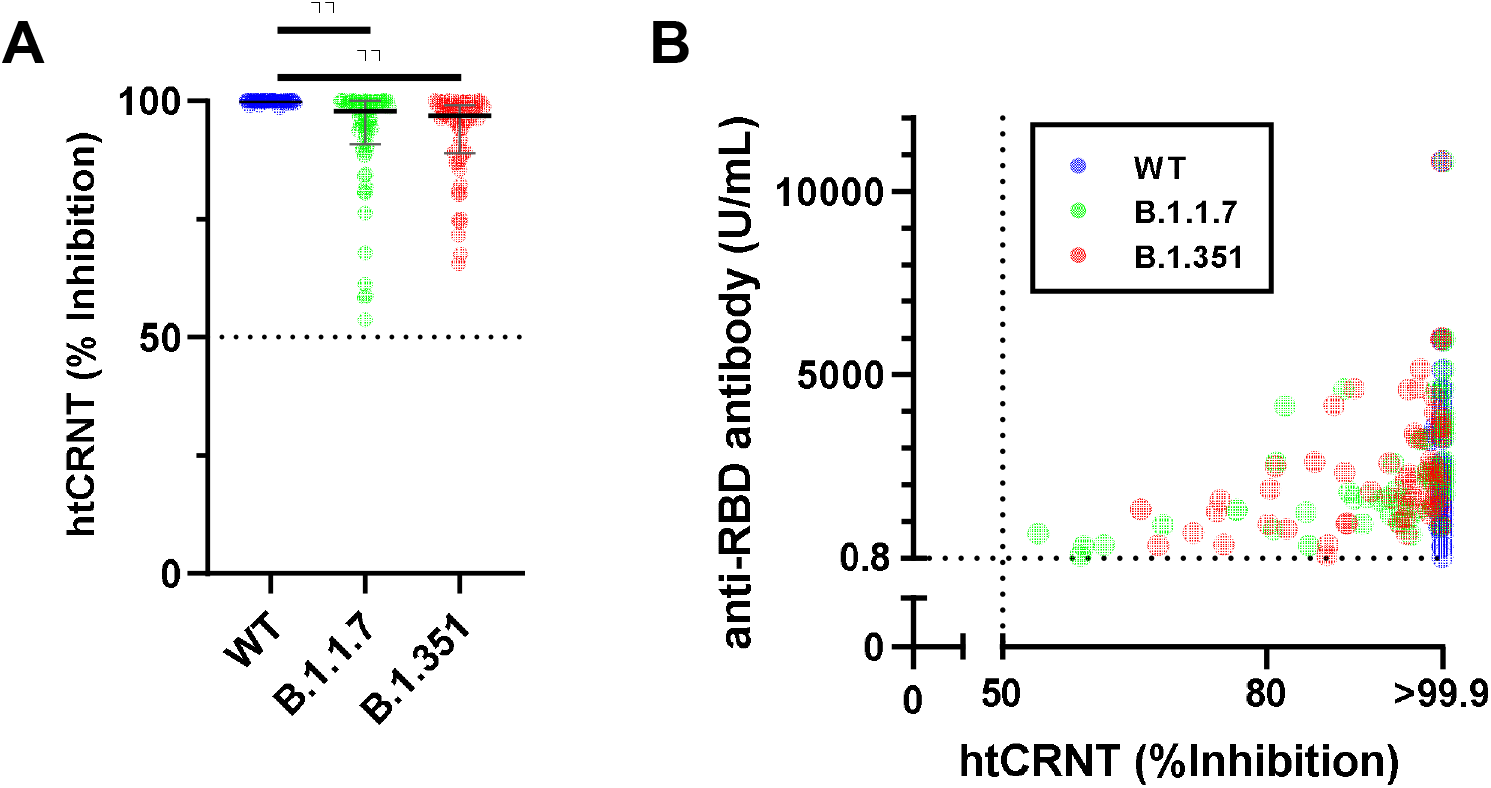
Cross-reaction against B.1.1.7 and B.1.351-derived variants in representative sera. (A) Neutralising activity against wild-type (WT), B.1.1.7-, and B.1.351-derived variants (n=61). (B) The relationship between neutralising activity and anti-RBD antibody levels. **, p < 0.01. Bars indicate medians with interquartile ranges.

### Relationship between vaccine-induced antibody and demographic characteristics

Finally, demographic characteristics were obtained from participants who answered the questionnaire and their test results were compared. A total of 237 participants provided their ages and symptoms after vaccination (**Table 1**). Of these, 21 had data with neutralisation against the variants.

**Table 1.**
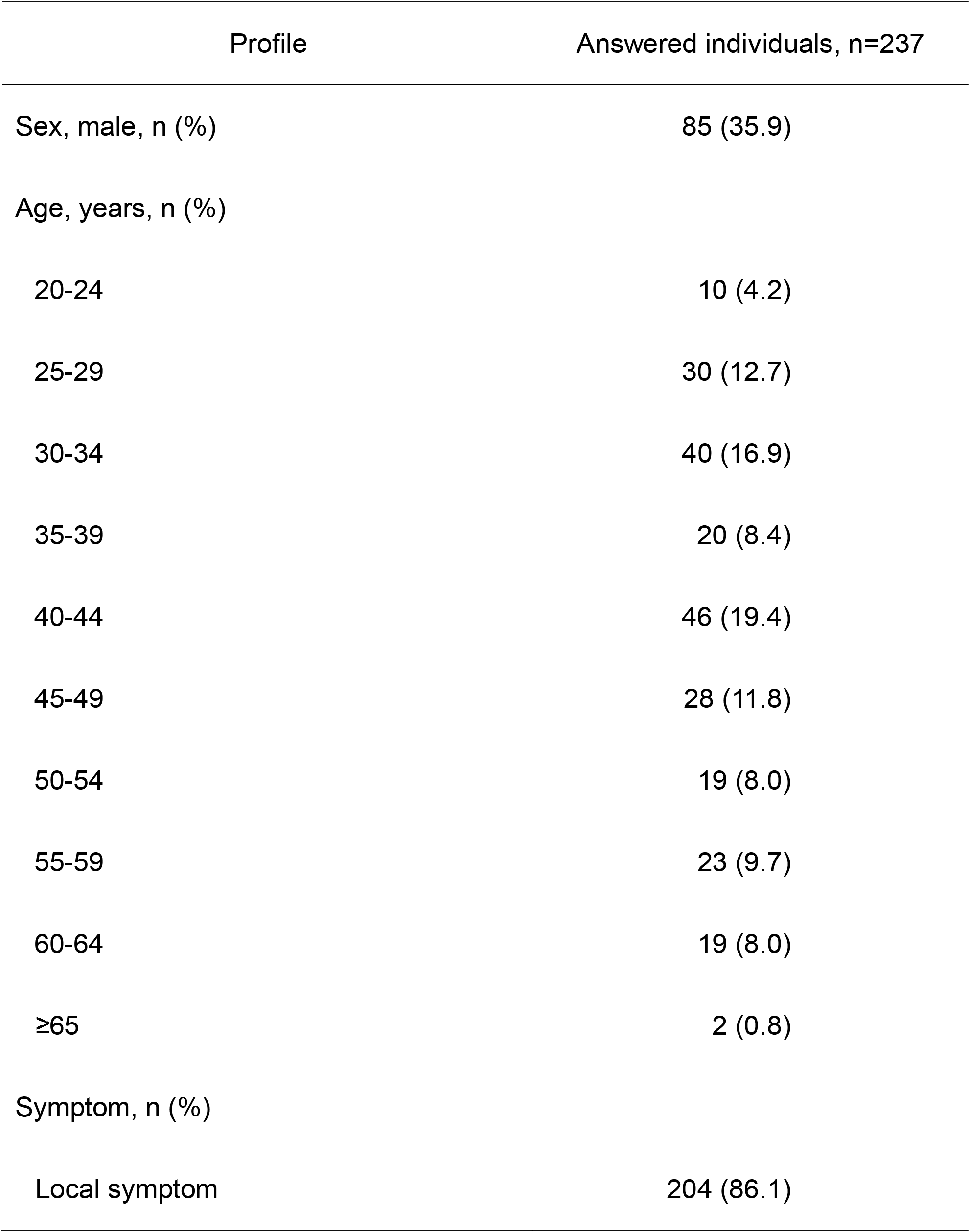
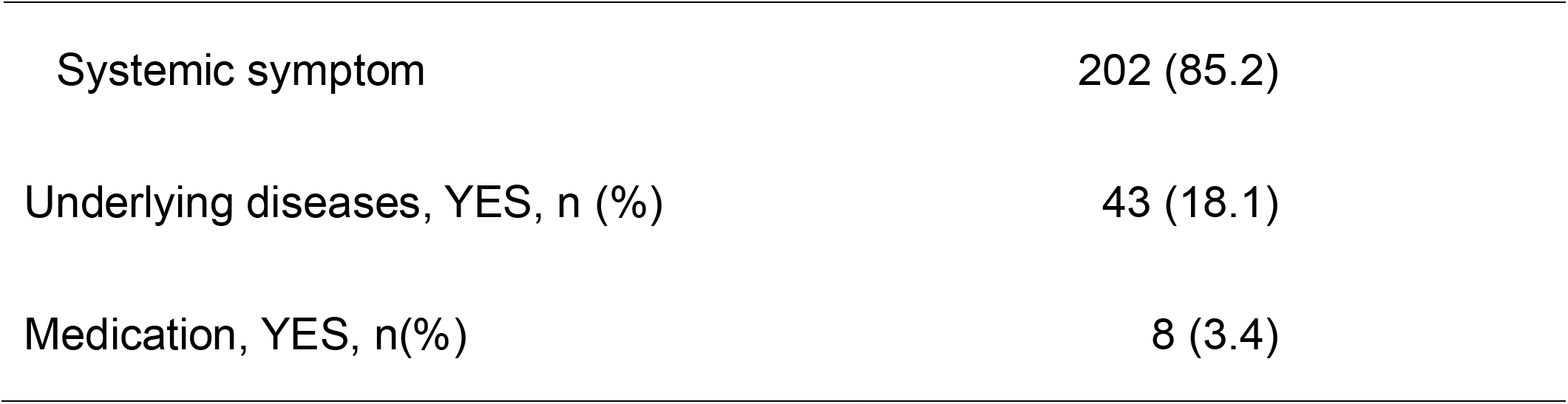
Demographic and clinical characteristics of the study participants who answered the questionnaire.

| Profile | Answered individuals, n=237 |
| --- | --- |
| Sex, male, n (%) | 85 (35.9) |
| Age, years, n (%) |  |
| 20-24 | 10 (4.2) |
| 25-29 | 30 (12.7) |
| 30-34 | 40 (16.9) |
| 35-39 | 20 (8.4) |
| 40-44 | 46 (19.4) |
| 45-49 | 28 (11.8) |
| 50-54 | 19 (8.0) |
| 55-59 | 23 (9.7) |
| 60-64 | 19 (8.0) |
| ≥65 | 2 (0.8) |
| Symptom, n (%) |  |
| Local symptom | 204 (86.1) |
| Systemic symptom | 202 (85.2) |
| Underlying diseases, YES, n (%) | 43 (18.1) |
| Medication, YES, n(%) | 8 (3.4) |

The values of the anti-RBD antibody test in females (median 2345, IQR 1388–3960) were significantly higher than those in males (median 1967, IQR 1086–2849; p<0.05) (**Fig 3A**) and were negatively correlated with age for females (r = -0.31, 95% CI -0.45 to -0.16; p<0.01) but not for males (r = -0.08; 95% CI -0.29 to 0.14; p=0.47) (**Fig 3B**). For the neutralisation, while the values of htCRNT against the wild-type were not correlated with age, those against the variants decreased in an age-dependent manner, with the B.1.351-derived variant showing significance (B.1.1.7-derived variant: p = 0.07, r = -0.40, 95% CI -0.71 to 0.03; B.1.351-derived variant, p < 0.01, r = -0.57, 95% CI -0.81 to 0.19) (**Fig 3C**).

**Figure 3.**
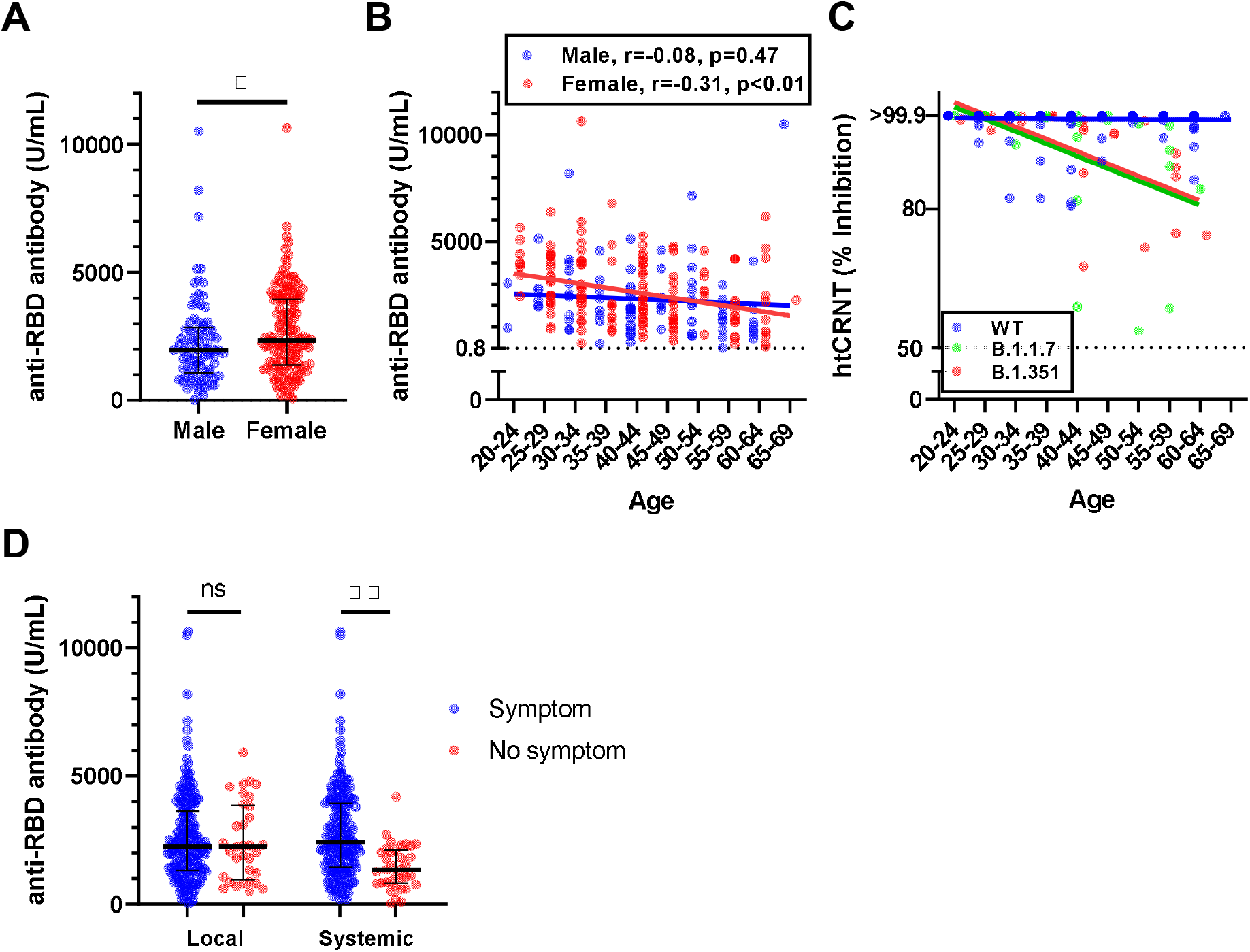
Relationship of the vaccine-induced antibody and demographic characteristics in questionnaire-answered population. (A) Anti-RBD antibody levels in males and females (n=237). (B) Relationship between anti-RBD antibody levels and age. (C) Relationship between htCRNT levels and age (for WT pseudotyped virus; n= 237, for B.1.1.7- and B.1.351-derived variants; n = 21). (D) Anti-RBD antibody levels and local or systemic symptoms. *; p < 0.05, **; p < 0.01, ns; not significance. Bars indicate medians with interquartile ranges. RBD, receptor-binding domain; htCRNT, high-throughput chemiluminescent reduction neutralising test.

The systemic symptoms after vaccination related to the anti-RBD antibody levels and the number of individuals without systemic symptoms significantly decreased compared with those with systemic symptoms (with systemic symptoms: median 2426, IQR 1450–3933 vs. without systemic symptoms: median 1347, IQR 818–2125; p<0.01) (**Fig 3D**). There was no significant change in local symptoms. For individuals with underlying diseases or medications, there were no significant differences in anti-RBD antibody levels.

## Discussion

After BNT162b2 vaccination, sufficient antibodies in terms of both quality and quantity were acquired. Surprisingly, compared with our previous findings in convalescent COVID-19 (CRNT, median 83.5, IQR 64.1–90.0; anti-RBD antibody test, median 35.0, IQR 7.63–137.0) [8], the amount of the anti-RBD antibody was 60.3-fold elevated and vaccinated sera neutralised pseudotyped viral infection almost completely. These findings suggest that the BNT162b2 vaccine can provide sufficient antibodies that surpass the immunity acquired by natural infection. In addition, individuals who have recovered from COVID-19 have generated some protective immunity; however, the vaccine can enhance their immune responses.

Similar to previous reports [4, 6], our findings support the notion that vaccinated sera can cross-react with SARS-CoV-2 variants. The reduction in neutralisation against the B.1.351 variant has been widely recognized [7, 10]; however, for the B.1.1.7 variant, some reports indicated that the neutralisation activity is similar to that of the wild-type [6, 11] and some reported a reduced level compared to the wild-type [4, 5, 12]. In the present study, although the neutralisation against variants was significantly reduced compared to the wild-type, almost all sera still had a >50% inhibitory effect. Because approximately half of the convalescent sera did not show >50% inhibition in our previous study, vaccine-induced antibodies are likely to be more effective against variants than infection-induced antibodies. In addition, efficacy against variants seems to be quantitatively related with a larger amount of the antibody induced by the vaccine.

Sex differences were observed, where females developed higher antibody levels in vaccine-induced immunity. Similar differences in vaccination have also been reported in previous reports [13, 14]. The acquisition of antibody levels was reduced in an age-dependent manner in females. Similarly, age-related reduction was indicated after BNT162b2 vaccination using another commercial antibody test [15]. These findings are consistent with a general understanding of vaccinated immunity [16]. An important aspect of the present study is that it shows sufficient immunity even for the oldest participants. However, because this study did not include elderly people, further examination will be necessary. Neutralisation against variants was maintained at >50% inhibition but showed age-dependent reduction. There was a significant difference only against the B.1.351-derived variant; however, it is reasonable to think that both the B.1.1.7- and B.1.351-derived variants tend to be similar. Meanwhile, it remains unknown how much a reduction in neutralisation will impact one’s future risk of COVID-19. Whether these findings in age and sex have clinical impact should be monitored in the future; however, cross-reaction to variants can support secondary immunity.

The limitations of the current study are as follows. The efficacy of SARS-CoV-2 vaccination among people whose immunity may be unstable, such as children, elderly people, and individuals with underlying diseases, is unknown. It is still unknown how long immunity is maintained after vaccination. Therefore, we plan to conduct a follow-up study for the participants. Lastly, it is impossible to exclude possible participants who had already acquired immunity because sera prior to vaccination were not assessed.

In conclusion, the BNT162b2 vaccine produced a sufficient level of antibodies with a neutralising effect against WT SARS-CoV-2 and emerging variants. Antibody induction can be affected by age and sex but is still induced to a sufficient level.

## Data Availability

All available data are in the manuscript.

## Acknowledgments

We thank all the staff at Toyama University Hospital for their help in collecting specimens and questionnaires. We would like to thank Mayu Somekawa for the arrangement of the specimens.

## Contribution

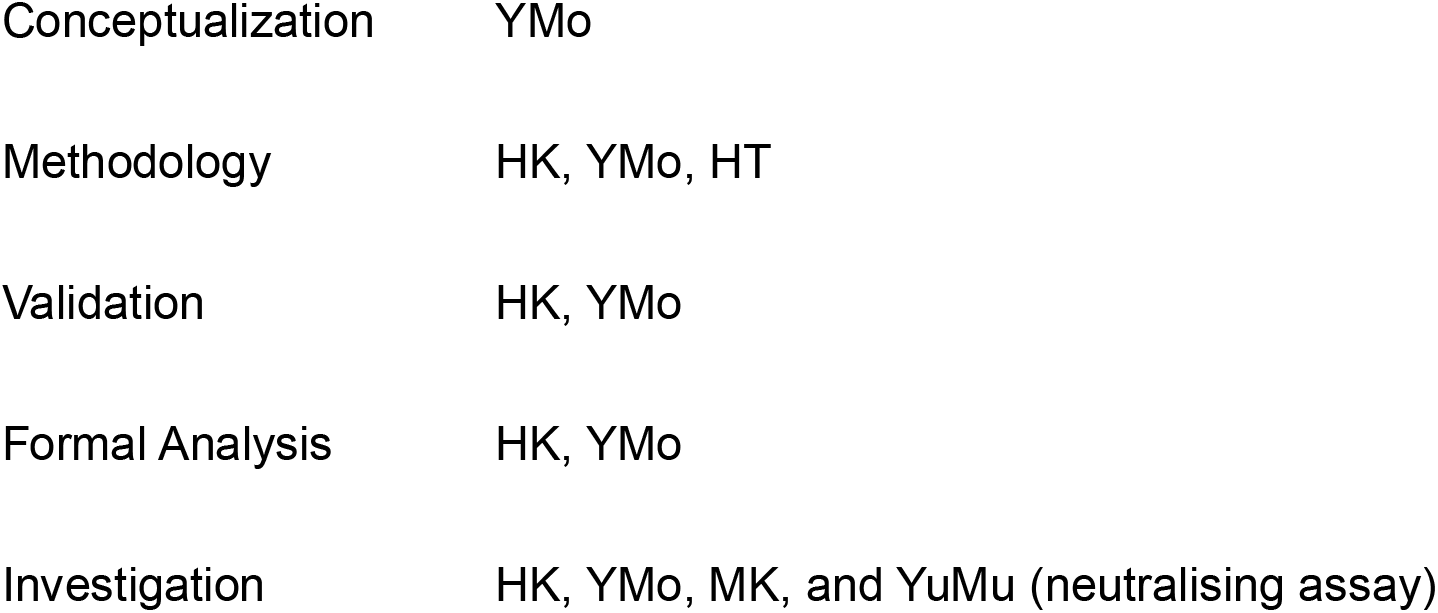
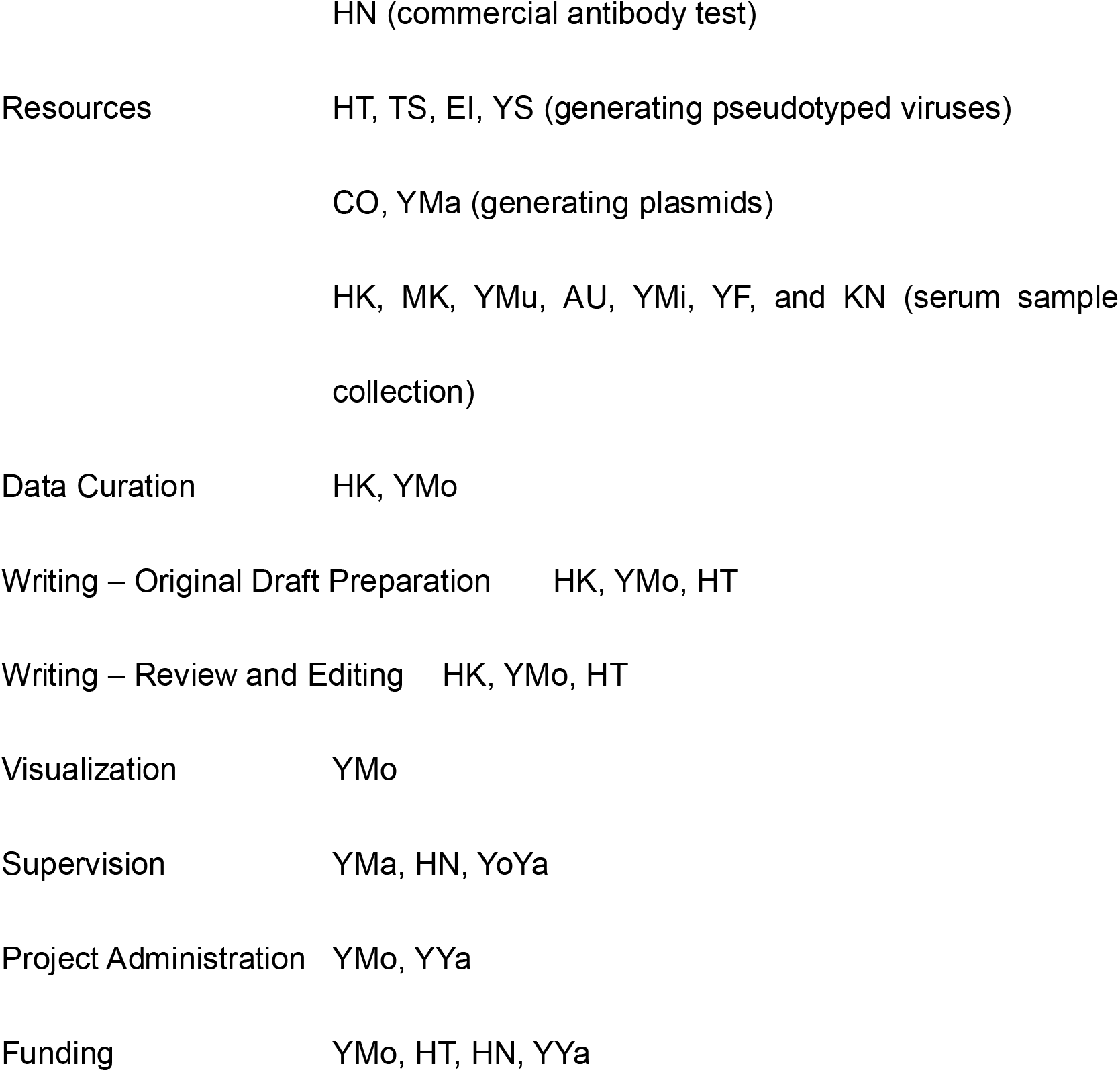

## References

[1] Polack FP, Thomas SJ, Kitchin N, Absalon J, Gurtman A, Lockhart S, Perez JL, Pérez Marc G, Moreira ED, Zerbini C, Bailey R, Swanson KA, Roychoudhury S, Koury K, Li P, Kalina WV, Cooper D, Frenck RW, Jr., Hammitt LL, Türeci Ö, Nell H, Schaefer A, Ünal S, Tresnan DB, Mather S, Dormitzer PR, Sahin U, Jansen KU, Gruber WC. Safety and Efficacy of the BNT162b2 mRNA Covid-19 Vaccine. New England J Med 2020; 383: 2603–2615.

[2] Starr TN, Greaney AJ, Hilton SK, Ellis D, Crawford KH, Dingens AS, Navarro MJ, Bowen JE, Tortorici MA, Walls AC. Deep mutational scanning of SARS-CoV-2 receptor binding domain reveals constraints on folding and ACE2 binding. Cell 2020; 182: 1295-1310.e20.

[3] Shrock E, Fujimura E, Kula T, Timms RT, Lee I-H, Leng Y, Robinson ML, Sie BM, Li MZ, Chen Y. Viral epitope profiling of COVID-19 patients reveals cross-reactivity and correlates of severity. Science 2020 in press.

[4] Xie X, Liu Y, Liu J, Zhang X, Zou J, Fontes-Garfias CR, Xia H, Swanson KA, Cutler M, Cooper D, Menachery VD, Weaver SC, Dormitzer PR, Shi PY. Neutralization of SARS-CoV-2 spike 69/70 deletion, E484K and N501Y variants by BNT162b2 vaccine-elicited sera. Nat Med 2021; 27: 620–621.

[5] Supasa P, Zhou D, Dejnirattisai W, Liu C, Mentzer AJ, Ginn HM, Zhao Y, Duyvesteyn HM, Nutalai R, Tuekprakhon A. Reduced neutralization of SARS-CoV-2 B. 1.1. 7 variant by convalescent and vaccine sera. Cell 2021 in press.

[6] Kuzmina A, Khalaila Y, Voloshin O, Keren-Naus A, Boehm-Cohen L, Raviv Y, Shemer-Avni Y, Rosenberg E, Taube R. SARS-CoV-2 spike variants exhibit differential infectivity and neutralization resistance to convalescent or post-vaccination sera. Cell Host Microb 2021; 29: 522-528.e2.

[7] Edara VV, Norwood C, Floyd K, Lai L, Davis-Gardner ME, Hudson WH, Mantus G, Nyhoff LE, Adelman MW, Fineman R, Patel S, Byram R, Gomes DN, Michael G, Abdullahi H, Beydoun N, Panganiban B, McNair N, Hellmeister K, Pitts J, Winters J, Kleinhenz J, Usher J, O’Keefe JB, Piantadosi A, Waggoner JJ, Babiker A, Stephens DS, Anderson EJ, Edupuganti S, Rouphael N, Ahmed R, Wrammert J, Suthar MS. Infection- and vaccine-induced antibody binding and neutralization of the B.1.351 SARS-CoV-2 variant. Cell Host Microb 2021; 29: 516-521.e3.

[8] Morinaga Y, Tani H, Terasaki Y, Nomura S, Kawasuji H, Shimada T, Igarashi E, Saga Y, Yoshida Y, Yasukochi R, Kaneda M, Murai Y, Ueno A, Miyajima Y, Fukui Y, Nagaoka K, Ono C, Matsuura Y, Fujimura T, Ishida Y, Oishi K, Yamamoto Y. Correlation of the commercial anti-SARS-CoV-2 receptor binding domain antibody test with the chemiluminescent reduction neutralizing test and possible detection of antibodies to emerging variants. medRxiv in preperation.

[9] Tani H, Kimura M, Tan L, Yoshida Y, Ozawa T, Kishi H, Fukushi S, Saijo M, Sano K, Suzuki T, Kawasuji H, Ueno A, Miyajima Y, Fukui Y, Sakamaki I, Yamamoto Y, Morinaga Y. Evaluation of SARS-CoV-2 neutralizing antibodies using a vesicular stomatitis virus possessing SARS-CoV-2 spike protein. Virology J 2021; 18: 16.

[10] Hoffmann M, Arora P, Groß R, Seidel A, Hörnich BF, Hahn AS, Krüger N, Graichen L, Hofmann-Winkler H, Kempf A, Winkler MS, Schulz S, Jäck HM, Jahrsdörfer B, Schrezenmeier H, Müller M, Kleger A, Münch J, Pöhlmann S. SARS-CoV-2 variants B.1.351 and P.1 escape from neutralizing antibodies. Cell 2021 in press.

[11] Planas D, Bruel T, Grzelak L, Guivel-Benhassine F, Staropoli I, Porrot F, Planchais C, Buchrieser J, Rajah MM, Bishop E, Albert M, Donati F, Prot M, Behillil S, Enouf V, Maquart M, Smati-Lafarge M, Varon E, Schortgen F, Yahyaoui L, Gonzalez M, De Sèze J, Péré H, Veyer D, Sève A, Simon-Lorière E, Fafi-Kremer S, Stefic K, Mouquet H, Hocqueloux L, van der Werf S, Prazuck T, Schwartz O. Sensitivity of infectious SARS-CoV-2 B.1.1.7 and B.1.351 variants to neutralizing antibodies. Nat Med 2021 in press.

[12] Bates TA, Leier HC, Lyski ZL, McBride SK, Coulter FJ, Weinstein JB, Goodman JR, Lu Z, Siegel SAR, Sullivan P, Strnad M, Brunton AE, Lee DX, Curlin ME, Messer WB, Tafesse FG. Neutralization of SARS-CoV-2 variants by convalescent and vaccinated serum. medRxiv 2021 DOI 10.1101/2021.04.04.21254881.

[13] Fischinger S, Boudreau CM, Butler AL, Streeck H, Alter G. Sex differences in vaccine-induced humoral immunity. Semin Immunopathol 2019; 41: 239–249.

[14] Fink AL, Engle K, Ursin RL, Tang WY, Klein SL. Biological sex affects vaccine efficacy and protection against influenza in mice. PNAS 2018; 115: 12477–12482.

[15] Müller L, Andrée M, Moskorz W, Drexler I, Walotka L, Grothmann R, Ptok J, Hillebrandt J, Ritchie A, Rabl D, Ostermann PN, Robitzsch R, Hauka S, Walker A, Menne C, Grutza R, Timm J, Adams O, Schaal H. Age-dependent immune response to the Biontech/Pfizer BNT162b2 COVID-19 vaccination. medRxiv 2021 DOI 10.1101/2021.03.03.21251066.

[16] Blomberg BB, Frasca D. Quantity, not quality, of antibody response decreased in the elderly. J Clin Invest 2011; 121: 2981–3.

